# Prospective validation of clinical deterioration predictive models prior to intensive care unit transfer among patients admitted to acute care cardiology wards

**DOI:** 10.1101/2023.12.18.23300152

**Authors:** Jessica Keim-Malpass, Liza P Moorman, J. Randall Moorman, Susan Hamil, Gholamreza Yousevfand, Oliver J Monfredi, Sarah J Ratcliffe, Katy N Krahn, Marieke K Jones, Matthew T Clark, Jamieson M Bourque

## Abstract

Very few predictive models have been externally validated in a prospective cohort following the implementation of an artificial intelligence analytic system. This type of real-world validation is critically important due to the risk of data drift, or changes in data definitions or clinical practices over time, that could impact model performance in contemporaneous real-world cohorts. In this work, we report the model performance of a predictive analytics tool that was developed prior to COVID-19 and demonstrates model performance during the COVID-19 pandemic. The analytic system (CoMET®, Nihon Kohden Digital Health Solutions LLC, Irvine, CA) was implemented in a randomized controlled trial that enrolled 10,422 patient visits in a 1:1 display-on display-off design. The CoMET scores were calculated for all patients but only displayed in the display-on arm. Only the control/display-off group is reported here because the scores could not alter care patterns. Of the 5184 visits in the display-off arm, 311 experienced clinical deterioration and care escalation, resulting in transfer to the intensive care unit (ICU), primarily due to respiratory distress. The model performance of CoMET was assessed based on areas under the receiver operating characteristic curve, which ranged from 0.732 to 0.745. The models were well-calibrated, and there were dynamic increases in the model scores in the hours preceding the clinical deterioration events. A hypothetical alerting strategy based on a rise in score and duration of the rise would have had good performance, with a positive predictive value more than 10-fold the event rate. We conclude that predictive statistical models developed five years before study initiation had good model performance despite the passage of time and the impact of the COVID-19 pandemic. We speculate that some of the model performance’s stability is due to continuous cardiorespiratory monitoring, which should not drift as practices, policies, and patient populations change.

**Clinical Trial registration:** ClinicalTrials.gov <u>NCT04359641</u>;

https://clinicaltrials.gov/ct2/show/NCT04359641.

## Introduction

While predictive analytic models have been advanced to the bedside, not many have been externally validated in a new data set, and far fewer have been implemented and used in clinical practice (Moorman 2021, Ruminski *et al* 2019, Keim-Malpass and Moorman 2021, de Hond *et al* 2022).

External validation in contemporaneous data is a particularly important aspect of the vision of predictive analytics monitoring at the bedside, as we know now that data can drift (Loftus *et al* 2022). That is to say, the nature and frequency of elements in the EHR change over time as practices and policies change, and new data that alters practice comes to light. (Finlayson *et al* 2021, Yang *et al* 2022)

For example, the myocardial band of creatine kinase was once the blood test of choice in patients suspected of acute myocardial infarction, but that has been replaced by troponin. Thus, a predictive model for heart attack that included the former and omitted the latter would be severely impaired.

Moreover, since no one model can fit all scenarios in the hospital, we typically train predictive models for specific targets and specific populations (Blackwell *et al* 2020). If the hospital unit patient population, acuity, or care patterns were to change its composition a great deal, as we saw during the COVID-19 pandemic, then these fine-tuned models might lose their accuracy.

We have argued in favor of the use of models based on cardiorespiratory physiology as a means to circumvent data drift. (Monfredi *et al* 2023) Our reasoning is that such observations as heart rate, respiratory rate, blood pressure, and the mathematical properties of continuous cardiorespiratory monitoring (*e*.*g*., physiological time series) are not subject to the ordering practices of clinicians and their association with clinical deterioration should be stable with respect to time.

As part of a recent randomized controlled trial of predictive analytics monitoring on consecutive patients admitted to an acute care cardiac medical-surgical ward, we calculated but did not display regression-based predictive analytics scores for over 4484 patients among 5184 visits randomized to the control arm. We sought to investigate the possibility that model performance changed because of changes in patient population or hospital practices over time. More than five years elapsed between model development and the randomized trial, and the trial started, by coincidence, at the beginning of the first wave of the pandemic in Charlottesville, VA. Here, we report the model performance of this predictive analytics tool. Since the data were not used for model development, this is a TRIPOD type IV prospective validation study (Collins *et al* 2015).

## Methods

Following Institutional Review Board approval from the University of Virginia, we undertook a 2-arm cluster randomized controlled trial to test the impact of a predictive analytics monitor display (CoMET®, Nihon Kohden Digital Health Solutions LLC, Irvine, CA.) on outcomes in consecutive patients admitted to acute care medical/cardiology, post-operative cardio-thoracic surgery, and medical/surgical hospital wards. The trial protocol has been published. (Keim-Malpass *et al* 2021) Briefly, we randomized clusters of four contiguous beds among the 80 beds spread over three wards. Clusters were re-randomized every two months. In the display-on arm, large computer monitor screens displayed the predicted risks of imminent cardiorespiratory events and of imminent cardiovascular events using CoMET. These models are logistic regression models with cubic splines based on physiological measurements from continuous cardiorespiratory monitoring data and on EHR elements of vital signs and laboratory tests. The models were trained separately on cardiorespiratory and cardiovascular events of clinical deterioration leading to escalation in care delivery. (Moss *et al* 2017, Ruminski *et al* 2019, Keim-Malpass *et al* 2022, Blackwell *et al* 2020)

Randomization began on January 4, 2021 and ended October 4, 2022. We enrolled 10,422 patient visits into the randomized controlled trial. The display-off arm comprised fewer than half of the sample and received usual care. This subgroup is the focus of this report on predictive model performance, as the scores were not available to clinicians to influence care and therefore potentially alter performance.

Clinical research coordinators prospectively followed enrolled patients and individually adjudicated transfers to the ICU. We analyzed only those transfers that were undertaken for true clinical deterioration as opposed to movement to the ICU for elective procedures. An independent extractor then reviewed all emergent ICU transfer cases to determine the type of clinical deterioration using earlier definitions (Blackwell *et al* 2020). Reasons for ICU transfer included: bleeding/acute hemorrhage, coronary artery disease (CAD), neurological disease (Neuro), arrhythmia, heart failure (HF), infection, and respiratory deterioration. Importantly, these categories are not mutually exclusive, so a patient could have multiple etiologies of clinical deterioration for a single ICU transfer event.

We tested the performance of the models as continuous risk predictors analyzed every 15 minutes. All analyses used the 12 hours preceding clinical deterioration requiring emergent ICU transfer as the event detection window, *i*.*e*., a positive binary classification was labeled a true positive if it occurred within 0 to 12 hours before emergent ICU transfer. We calculated (1) model discrimination using area under the receiver operating characteristic (ROC) curve (2) model calibration of predicted relative risk to observed relative risk, (3) dynamic changes in model outputs prior to the deterioration event with significance testing using a Wilcoxon sign rank test, (4) model binary classification performance with an alert for a specified amount and duration of model rise, and (5) empirical risk of emergent ICU transfer as a function of the joint distribution of estimated cardiovascular and cardiorespiratory risk.

We additionally estimated the observed risk of emergent ICU transfer in the next 12 hours as a function of the empirical distribution of cardiovascular and cardiorespiratory risk estimates. Using kernel density estimation, we calculated the two-dimensional density of risk estimates for all patients all the time, and also for data within 12 hours before emergent ICU transfer (*i*.*e*., event data). We estimated the kernel bandwidth using the event data and used the same bandwidth for event data and all data density estimates. The relative risk of ICU transfer, then, is the ratio of the event data density divided by the density of all data.

## Results

### Patient population

Over 22 months, there were 5184 visits among the 4482 patients. Table 1 shows the characteristics of the display-off arm patients at the time of entry into the study. The sample was predominantly male (58%) and white (77%) with a mean age at admission of 66 years old. Of these display-off arm patients, 311 patients (6.9%) emergently transferred to the ICU. We analyzed model outputs in 2,174,117 15-minute epochs, of which 0.59% were within 12 hours prior to an emergent ICU transfer.

**Table 1:** Admission Characteristics. * n(%) or median (IQR) are shown

| Characteristic | N = 4482 (%) |
| --- | --- |
| Sex |  |
| Female | 1875 (41.8) |
| Male | 2607 (58.2) |
| Race |  |
| White | 3470 (77.4) |
| Black | 787 (17.6) |
| Asian | 42 (0.9) |
| Other | 172 (3.8) |
| Unknown | 11 (0.2) |
| Ethnicity |  |
| Hispanic | 135 (3.0) |
| Non-Hispanic | 4333 (96.7) |
| Refused / Unknown | 14 (0.3) |
| Age at Admission | 66.0 (55.0, 75.0) |
| Hospital Unit |  |
| Medical-Cardiology | 1661 (37.1) |
| Cardio-Thoracic Surgery | 1508 (33.6) |
| Medical/Surgical | 1313 (29.3) |

\* n(%) or median (IQR) are shown

### Clinical events

Figure 1 shows the etiologies of clinical deterioration, which were often multiple, leading to emergent intensive care unit transfer. The UpSet plot represents overlapping attributes; for example, the overlapping set on the farthest right-hand column represents the number of patients transferred emergently to the ICU for arrhythmia, infection, and respiratory decompensation. The most common reason patients experienced clinical deterioration was due to respiratory distress (36.3%). This plot shows the wide diversity of reasons for decompensation along with various co-occurring etiologies.

**Figure 1.**
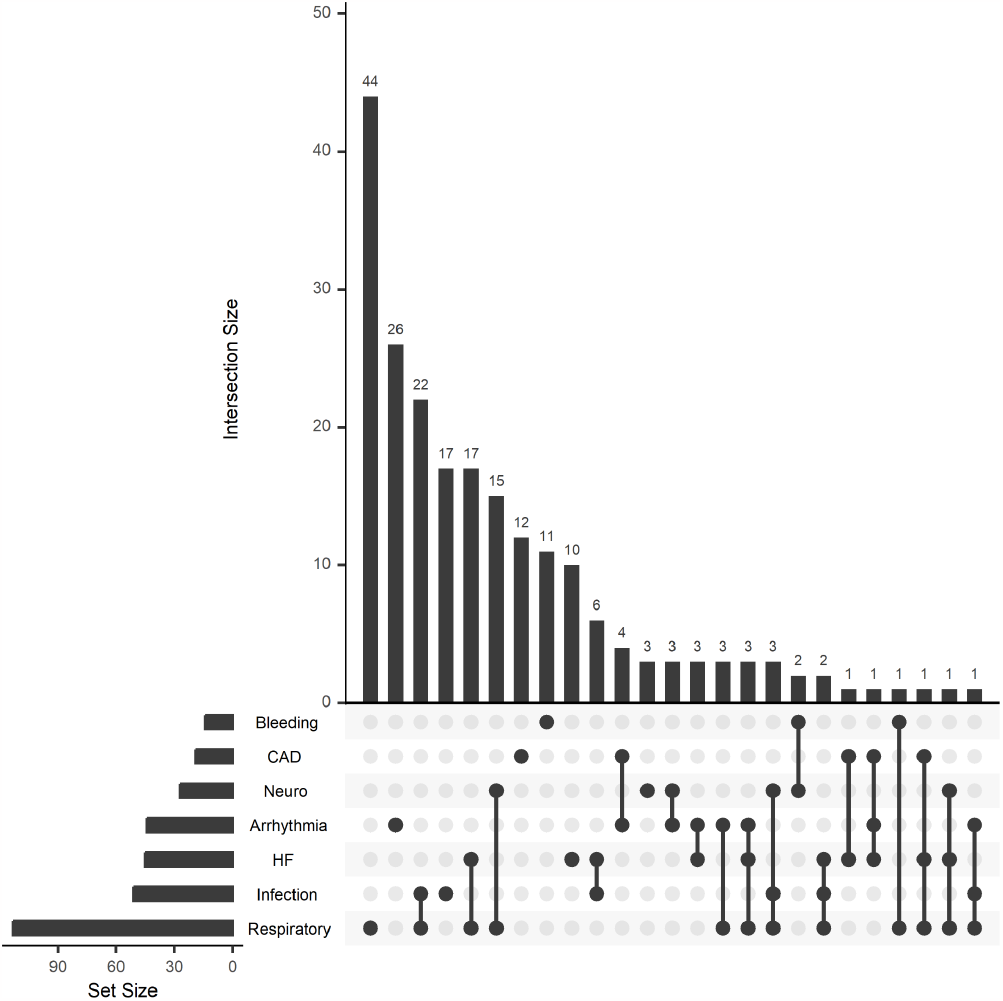
UpSet plot of additive reasons for emergent ICU transfer. The set represents overlapping attributes, for example, the first set on the right hand column represents the number of patients who had simultaneous arrhythmia, infection, and respiratory clinical deterioration events.

### Model performance: continuous risk prediction

We evaluated the discrimination of the models by measuring the area under the receiver-operating characteristic curve (AUC) on every 15-minute predicted risk. The cardiovascular model predictive performance was AUC = 0.732 (95% CI 0.701 - 0.762), the cardiorespiratory model predictive performance was AUC = 0.745 (95% CI 0.720 - 0.768), and the all-cause ICU transfer model predictive performance was AUC = 0.741 (95% CI 0.713 - 0.772). For reference, the all-cause ICU transfer model had an AUC of 0.729 in the training dataset (Moss *et al* 2017).

Figure 2 shows the calibration of the models. The major finding is that the points lie close to the dotted line of identity; the predicted risks were close to the observed risks. Thus, the model is calibrated well, though the respiratory model slightly underestimated the observed risk at the extreme high end.

**Figure 2a and 2b.**
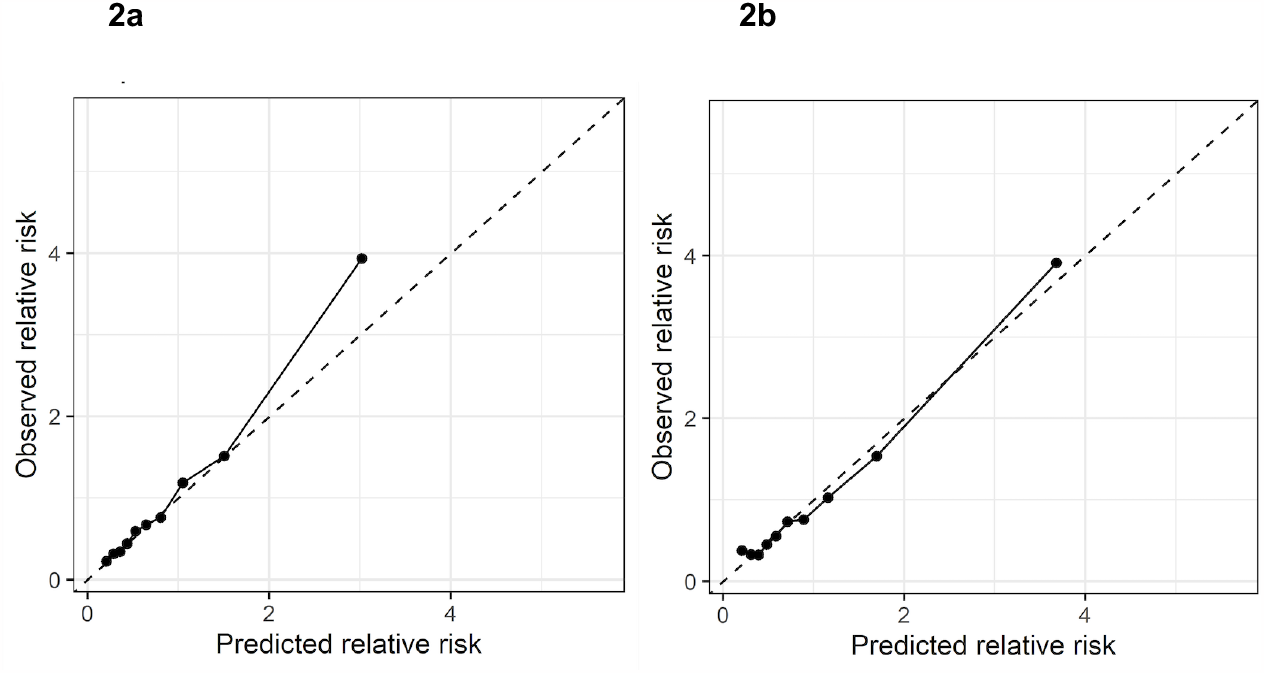
Calibration of the predictive models. 2a is the cardiorespiratory model, 2b cardiovascular model. The data points are the average observed risk plotted as a function of each decile of the CoMET score, *i*.*e*., predicted relative risk. Perfect calibration is shown as a dashed line of identity.

Figure 3 shows the diagnostic likelihood ratio, or the ratio of post-test risk to pre-test risk, of ICU transfer as a function of the range of the CoMET score. These ranges provide risk groups based on natural thresholds interpretable to end users. The likelihood ratio increases with increasing risk group and the risk is much higher in patients with scores >4.

**Figure 3a and 3b.**
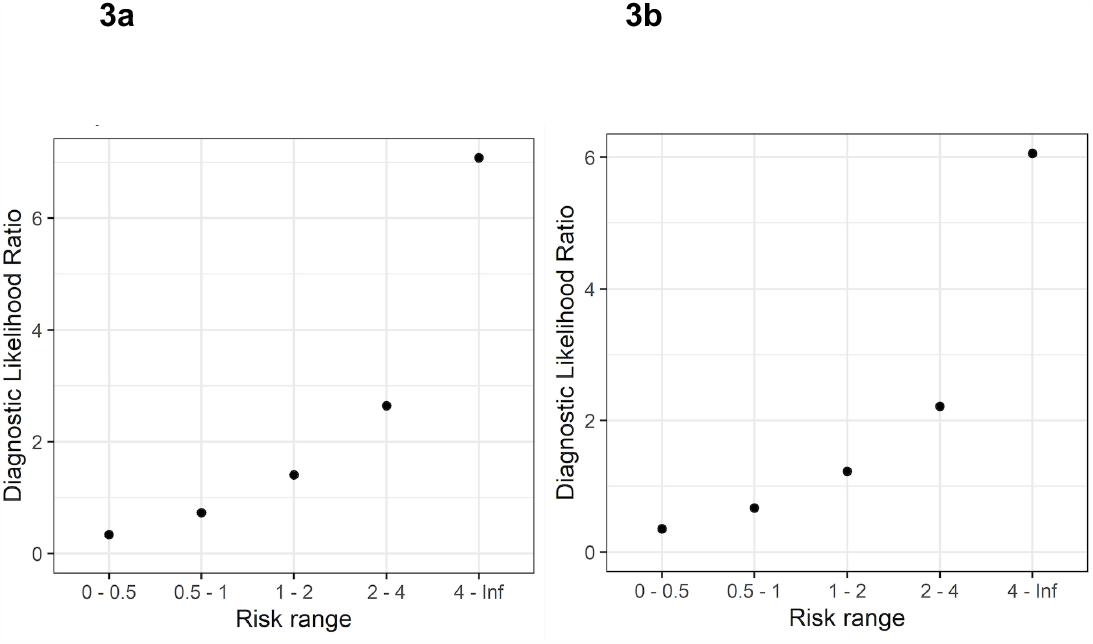
Calibration of the risk models (3a is the cardiorespiratory model, 3b cardiovascular model) by risk group. Each bin is inclusive at the lower bound and exclusive at the upper bound. This figure demonstrates the likelihood ratio of the event as a function of the CoMET scores. As expected, the likelihood of an event increases substantially as the model-predicted risk rises.

### Model performance: Dynamic changes before events

Figure 4 shows the average model scores as a function of time until the ICU transfer at time zero. The average relative risk rose from approximately 2-fold in the 48 to 24 hours prior to emergent ICU transfer to approximately 3-fold at transfer. Open circles denote a statistically significant (p < 0.05) rise in the CoMET score relative to scores for the same patients to 24 hours prior, and the average score begins to rise appreciably up to 8 hours before the event.

**Figure 4a and 4b.**
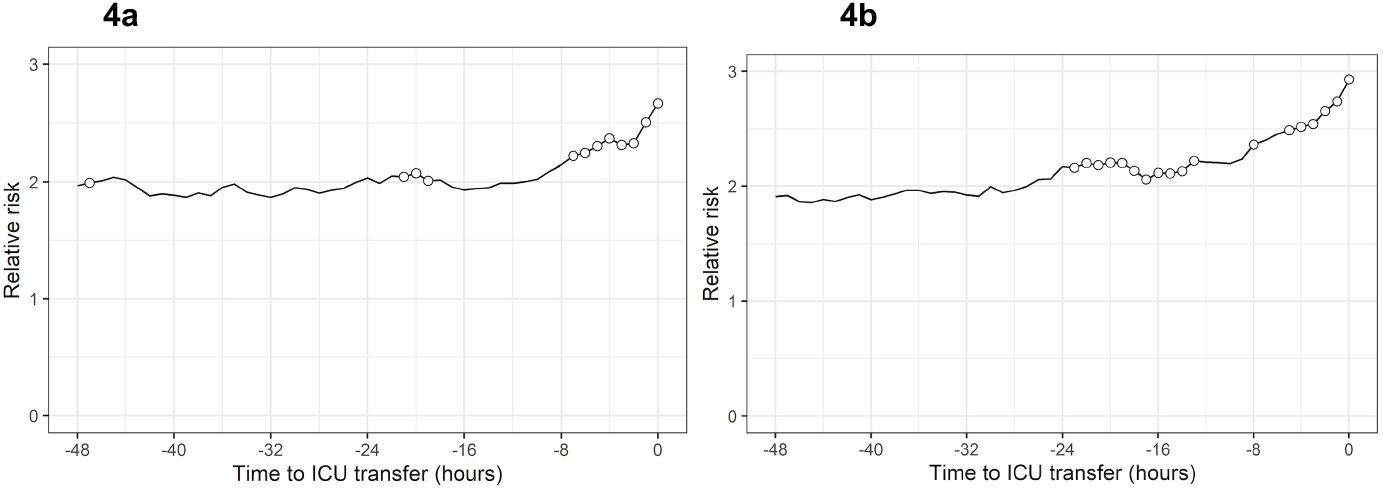
4a is the cardiorespiratory model, 4b cardiovascular model. The relative risk of an event rises as the time to ICU transfer nears. Open circles denote statistically significant increases in the score relative to 24 hours prior using a Wilcoxon sign rank test and 0.05 significance level.

### Model performance: binary classification

Here we defined hypothetical alerts based on the trend of an increase in score that remains high for a period of time. To calculate this, we averaged the model scores hourly and calculated the delta at each hour relative to the value 2 hours prior (each delta is therefore based on a 3-hour retrospective window). An alert is identified when an hourly score results in a delta that is greater than *d* and the average over the next *N* hours remains >90% of the hourly score. If the score rises from, for example, 1 to 4, this satisfies a delta *d* = 3. *N* is the number of hours for which the (running) average of the hourly scores is >=3.6 (90% of 4).

Figure 5 shows the positive predictive value (PPV) of an emergent ICU transfer as a function of the alert threshold. The PPV is shown relative to the event rate of 0.59%. Note that, unlike previous studies of alert strategies, only emergent ICU transfers were used to identify true positive alerts (Keim-Malpass *et al* 2020). The best binary classification performance occurs when an alert is issued for an increase in the CoMET score by 3 or 4 units followed by an elevated score for a period of 3 to 6 hours.

**Figure 5a and 5b.**
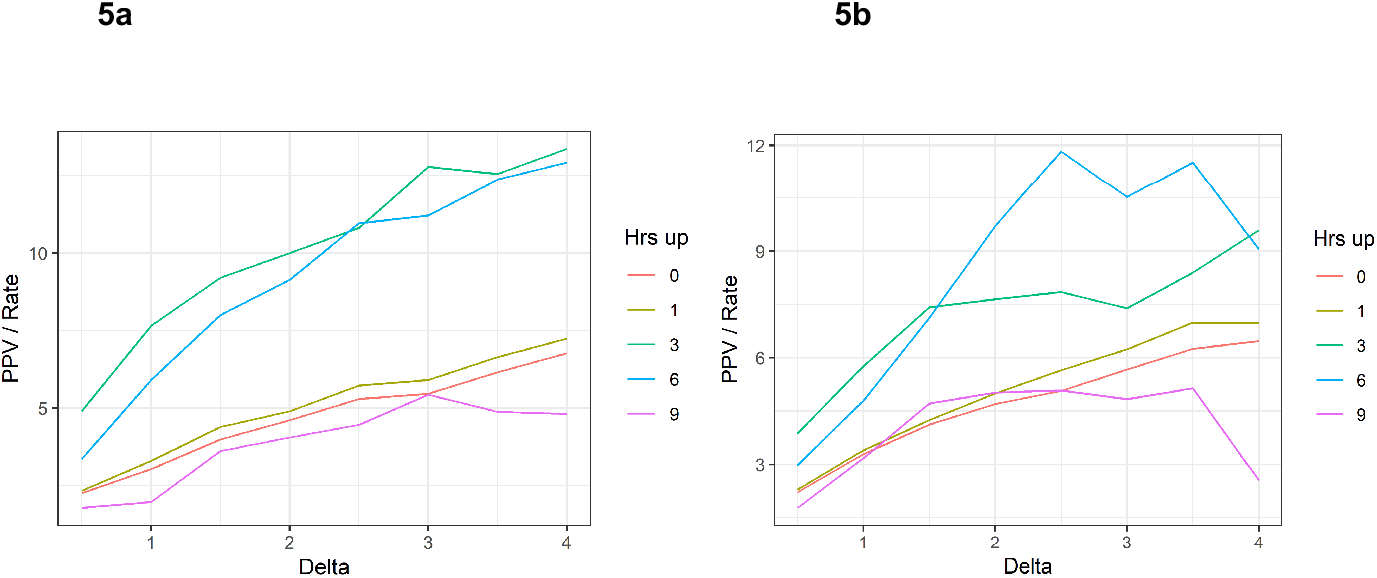
5a is the cardiorespiratory model, 5b cardiovascular model Evaluation as a trend-based thresholded alert demonstrated as the positive predictive value of both the rise and duration of risk score elevation. An alert is identified when an hourly score results in a delta that is greater than *d* and the average over the next *N* hours remains >90% of the hourly score. If the score rises from, for example, 1 to 4, this satisfies a delta *d* = 3. *N* is the number of hours for which the (running) average of the hourly scores is >=3.6 (90% of 4).

### Empirical risk of emergent ICU transfer

In Figure 6, grayscale quantifies the relative risk of emergent ICU transfer in the next 12 hours as a function of estimated cardiovascular risk (x-axis) and cardiorespiratory risk (y-axis). Whiter indicates higher risk. While Figure 2 shows the calibration of each model individually, that calibration in Figures 2A and 2B is the marginal distribution that would be obtained by averaging over the y- and x-axes of Figure 6, respectively. Figure 6 shows that the combination of 2 model risk estimates adds information about patient risk: while a patient with 2-fold cardiovascular risk has on average 2-fold risk of ICU transfer, that risk is less than one given a cardiorespiratory risk near zero, but greater than 3 given a cardiorespiratory risk near 5.

**Figure 6:**
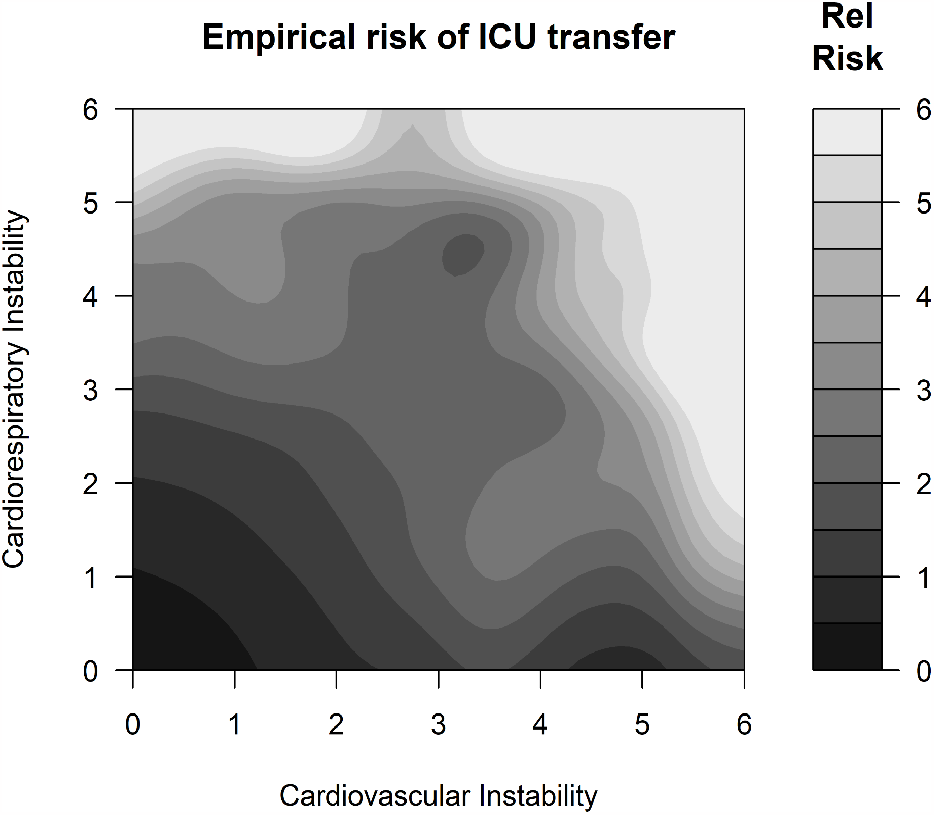
Empirical risk of emergent ICU transfer as a function of both estimated cardiovascular (x-axis) and cardiorespiratory risk (y-axis). Contour lines are lines of iso-risk. Color indicates relative risk at a given location (x, y). Black is low risk and white is high risk.

## Discussion

We studied the performance of two predictive risk models for clinical deterioration that were implemented in patients admitted to acute care cardiology, cardio-thoracic surgery, and medical-surgical wards in the context of a larger randomized controlled trial. Overall, the model performance and calibration were strong, and the score appropriately and dynamically rose in the 8 to 16 hours preceding clinical deterioration requiring ICU transfer. In addition to traditional categorical predictors, these models utilize predictors that are on a continuous scale and include mathematical representations derived from continuous cardiorespiratory monitoring data. The calibration can be compared favorably to the performance of heart rate characteristics for predicting neonatal sepsis, which also fits well at low and medium risk but, unlike these CoMET scores, over-estimated risk at the high end. (Lake *et al* 2014) The addition of continuous metrics can provide a more complete representation of patient physiological state, fills in the gaps between blood draws and nursing assessments, and represents a substantial advancement in dynamic predictive risk estimation in real-world settings (Monfredi *et al* 2021, Moss *et al* 2017).

Using contemporary events to study performance provides added confirmation of the robustness of these predictive models. While the models were trained using preceding retrospective data, this randomized controlled trial and subgroup study occurred during the COVID-19 pandemic. The predictive models used in this study and trained on earlier data continued to produce calibrated and accurate results regarding the risk of clinical deterioration despite substantial changes in hospital admission practices, fewer elective admissions for cardiac surgery, fewer emergency room visits for cardiac symptoms, and high turnover rates among nursing staff. This analysis is limited to the patients in the control (display-off arm) of the randomized controlled trial to evaluate model accuracy without contamination by potential changes in clinical practice based on the model results. More work is needed to understand the link between model rise and actual clinical action in real-world contexts.

We note that the cardiovascular model performed less well than the cardiorespiratory model based on AUC, though still commensurate with performance in the training data. We have consistently found higher predictive accuracy as represented by model AUC for models targeting urgent unplanned intubation for acute respiratory failure than for acute coronary events or stroke. One interpretation is that, on an acute cardiology ward, there are frequent episodes of cardiovascular decompensation that are treated without requiring intensive care that would count as false positives when evaluating the cardiovascular model, while respiratory decompensation that elevates the cardiorespiratory model more often requires escalation to the ICU. More detailed analysis of clinical action associated with presumed false positives may therefore elucidate the relationship between increased estimated risk and therapeutic interventions by the clinical team.

Here we evaluate the performance of two models targeted for different types of clinical deterioration: cardiovascular and cardiorespiratory. These models were designed to present a more accurate clinical context about the physiological system driving the deterioration. Our results indicate that risk estimates from each model individually are well-calibrated and dynamically increased prior to the event, but also that combining the two models provided better discrimination of patient risk. In addition to providing clinical detail about the mechanism of potential deterioration, then, for a given estimate from one model the estimate from the other model further refines patient risk. This observation provides further evidence that models should be tailored to specific patient populations and events.

This work also demonstrated that an alerting strategy based on both a rise in score and a continued elevation over a duration of several hours promises good performance, with PPV more than 10-fold increased over the event rate: a random classifier would have PPV equal to the event rate. We have evaluated these and other models for their performance as a basis for alerting clinicians of patient deterioration. Rather than use a single observed value, we have used the pattern of a large, abrupt spike in the risk score. In this way, we have found positive predictive values (PPV) ranging from 14% to nearly 50%. (Sullivan *et al* 2014, Keim-Malpass *et al* 2020) In a retrospective analysis, the models reported here have a PPV of 24% for a combined endpoint of unplanned ICU transfer (the subject of this report), infection workup, rapid response team visit, myocardial infarction, stroke, unplanned surgery, initiation of cardiopulmonary resuscitation, and death. (Keim-Malpass *et al* 2020)

While any rise in estimated risk must be due to some derangement in one or more physiological parameters in the model, a rise that resolves quickly points to clinical or self-resolution of said derangement while a rise that remains elevated may represent unrecognized derangement or one resistant to treatment. The rise in risk score by duration, representing a cumulative impact to the severity of illness, therefore offers a novel approach to predictive analytics monitoring in real-world use. This points to the need for additional study focusing on person-centered modeling approaches that rely on the change from a patient’s own baseline risk (Keim-Malpass *et al* 2020, Kausch *et al* 2022, Keim-Malpass and Kausch 2023)

We conclude that signatures of cardiorespiratory and cardiovascular illness are present in the continuous cardiorespiratory monitoring, vital signs, and laboratory data in patients hospitalized in an acute care cardiology ward. These statistical predictive models, developed five years previously, had good predictive performance despite the passage of time and the impact of the COVID-19 pandemic. We attribute some of the model performance’s stability to continuous cardiorespiratory monitoring, which should not drift as practices, policies, and patient populations change. Additional research on model impact on clinical actions, additional alert-based strategies, and patient-centered modeling approaches will provide added understanding in this important field.

## Data availability

All data produced in the present study are available upon reasonable request to the authors

## Bibliography

Blackwell J N, Keim-Malpass J, Clark M T, Kowalski R L, Najjar S N, Bourque J M, Lake D E and Moorman J R 2020 Early Detection of In-Patient Deterioration: One Prediction Model Does Not Fit All. Crit. Care Explor. 2 e0116

Collins G S, Reitsma J B, Altman D G, Moons K G M and TRIPOD Group 2015 Transparent reporting of a multivariable prediction model for individual prognosis or diagnosis (TRIPOD): the TRIPOD statement. The TRIPOD Group. Circulation 131 211–9

Finlayson S G, Subbaswamy A, Singh K, Bowers J, Kupke A, Zittrain J, Kohane I S and Saria S 2021 The clinician and dataset shift in artificial intelligence. N. Engl. J. Med. 385 283–6

de Hond A A H, Leeuwenberg A M, Hooft L, Kant I M J, Nijman S W J, van Os H J A, Aardoom J J, Debray T P A, Schuit E, van Smeden M, Reitsma J B, Steyerberg E W, Chavannes N H and Moons K G M 2022 Guidelines and quality criteria for artificial intelligence-based prediction models in healthcare: a scoping review. npj Digital Med. 5 2

Kausch S L, Sullivan B, Spaeder M C and Keim-Malpass J 2022 Individual illness dynamics: An analysis of children with sepsis admitted to the pediatric intensive care unit PLOS Digit Health 1 e0000019

Keim-Malpass J, Clark M T, Lake D E and Moorman J R 2020 Towards development of alert thresholds for clinical deterioration using continuous predictive analytics monitoring. J. Clin. Monit. Comput. 34 797–804

Keim-Malpass J and Kausch S L 2023 Data Science and Precision Oncology Nursing: Creating an Analytic Ecosystem to Support Personalized Supportive Care across the Trajectory of Illness. Semin. Oncol. Nurs. 151432

Keim-Malpass J and Moorman L P 2021 Nursing and precision predictive analytics monitoring in the acute and intensive care setting: An emerging role for responding to COVID-19 and beyond. International Journal of Nursing Studies Advances 3 100019

Keim-Malpass J, Ratcliffe S J, Moorman L P, Clark M T, Krahn K N, Monfredi O J, Hamil S, Yousefvand G, Moorman J R and Bourque J M 2021 Predictive Monitoring-Impact in Acute Care Cardiology Trial (PM-IMPACCT): Protocol for a Randomized Controlled Trial. JMIR Res. Protoc. 10 e29631

Keim-Malpass J, Moorman L P, Monfredi O J, Clark M T and Bourque J M 2022 Beyond prediction: Off-target uses of artificial intelligence-based predictive analytics in a learning health system Learn. Health Sys.

Lake D E, Fairchild K D and Moorman J R 2014 Complex signals bioinformatics: evaluation of heart rate characteristics monitoring as a novel risk marker for neonatal sepsis. J. Clin. Monit. Comput. 28 329–39

Loftus T J, Tighe P J, Ozrazgat-Baslanti T, Davis J P, Ruppert M M, Ren Y, Shickel B, Kamaleswaran R, Hogan W R, Moorman J R, Upchurch G R, Rashidi P and Bihorac A 2022 Ideal algorithms in healthcare: Explainable, dynamic, precise, autonomous, fair, and reproducible PLOS Digit Health 1 e0000006

Monfredi O, Keim-Malpass J and Moorman J R 2021 Continuous cardiorespiratory monitoring is a dominant source of predictive signal in machine learning for risk stratification and clinical decision support. Physiol. Meas. 42

Monfredi O J, Moore C C, Sullivan B A, Keim-Malpass J, Fairchild K D, Loftus T J, Bihorac A, Krahn K N, Dubrawski A, Lake D E, Moorman J R and Clermont G 2023 Continuous ECG monitoring should be the heart of bedside AI-based predictive analytics monitoring for early detection of clinical deterioration. J. Electrocardiol. 76 35–8

Moorman L P 2021 Principles for real-world implementation of bedside predictive analytics monitoring. Appl. Clin. Inform. 12 888–96

Moss T J, Clark M T, Calland J F, Enfield K B, Voss J D, Lake D E and Moorman J R 2017 Cardiorespiratory dynamics measured from continuous ECG monitoring improves detection of deterioration in acute care patients: A retrospective cohort study. PLoS ONE 12 e0181448

Ruminski C M, Clark M T, Lake D E, Kitzmiller R R, Keim-Malpass J, Robertson M P, Simons T R, Moorman J R and Calland J F 2019 Impact of predictive analytics based on continuous cardiorespiratory monitoring in a surgical and trauma intensive care unit. J. Clin. Monit. Comput. 33 703–11

Sullivan B A, Grice S M, Lake D E, Moorman J R and Fairchild K D 2014 Infection and other clinical correlates of abnormal heart rate characteristics in preterm infants. J. Pediatr. 164 775–80

Yang J, Karstens L, Ross C and Yala A 2022 AI Gone Astray: Technical Supplement arXiv

